## Supplementary figures and images for "Longitudinal analysis at three oral sites links oral microbiota to clinical outcomes in allogeneic hematopoietic stem-cell transplant"

### Additional file 1: Timelines of antibiotic usage

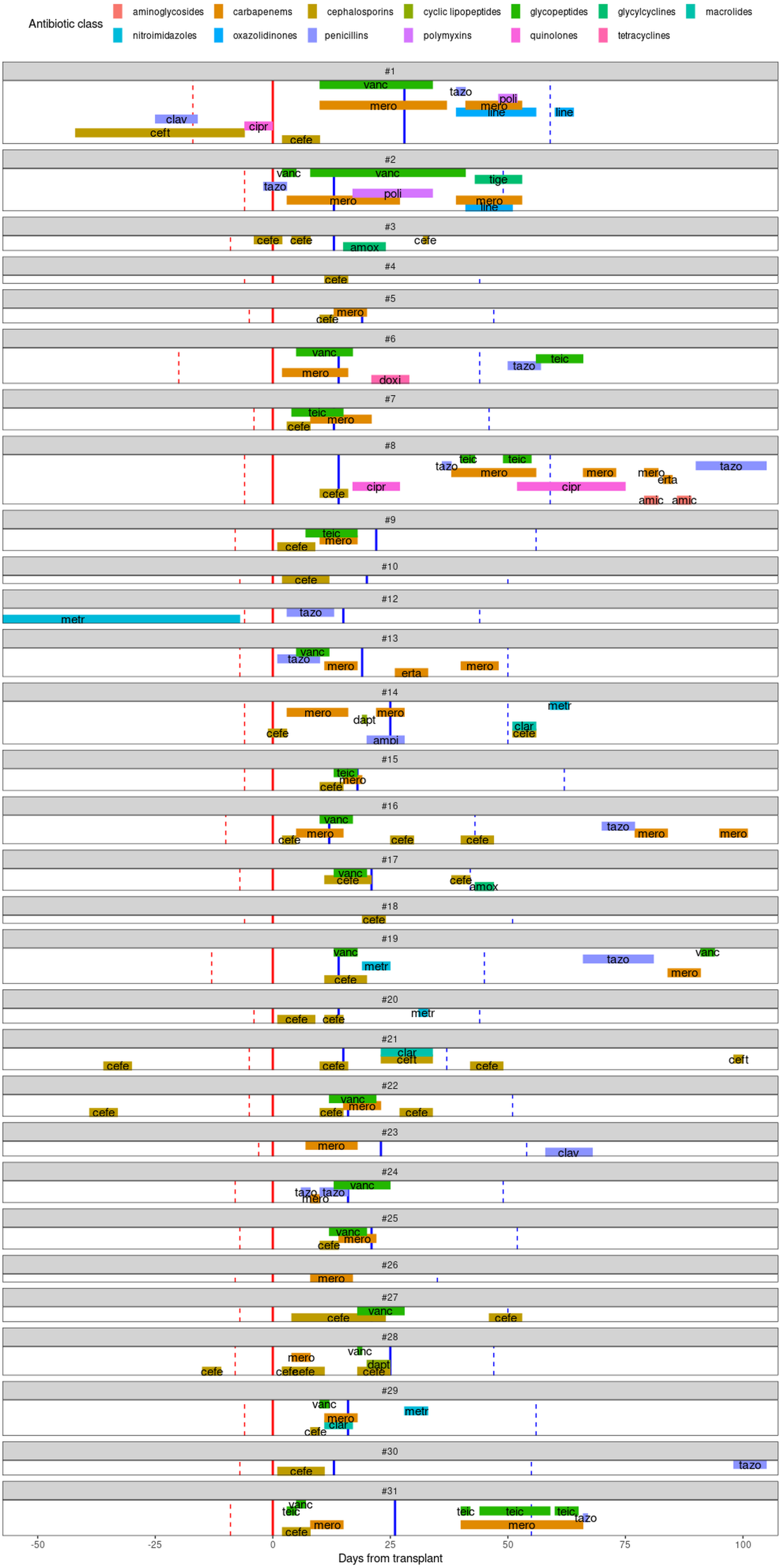
