## Additional file 2: Supplementary tables and figures for "Longitudinal analysis at three oral sites links oral microbiota to clinical outcomes in allogeneic hematopoietic stem-cell transplant"

**Table S1**

| | **n = 31** |
|---|---|
| **Age in years (median, range)** | 50 (19–73) |
| **Sex (male)** | 17 (55%) |
| **Underlying disease** | |
| Acute leukemia | 18 (58%) |
| Myeloid | 11 (35%) |
| Lymphocytic | 7 (23%) |
| Other | 13 (42%) |
| Non-Hodgkin lymphoma | 5 (16%) |
| Myelodysplastic syndrome | 5 (16%) |
| Chronic myeloid leukemia | 1 (3%) |
| Chronic lymphocytic leukemia | 1 (3%) |
| Multiple myeloma | 1 (3%) |
| **HCT-CI** | |
| 0 | 16 (52%) |
| 1-2 | 9 (29%) |
| ≥3 | 6 (19%) |
| **DRI** | |
| Low-intermediate | 18 (58%) |
| High | 13 (42%) |
| **Conditioning intensity** | |
| Reduced intensity | 19 (61%) |
| Myeloablative | 12 (39%) |
| **Total body irradiation** | 11 (35%) |
| **T-cell depletion** | 16 (52%) |
| **Graft source** | |
| Bone marrow | 10 (32%) |
| Peripheral blood | 21 (68%) |
| **Donor** | |

| | |
|---|---|
| MRD | 9 (29%) |
| Haploidentical | 10 (32%) |
| MUD/MMUD | 12 (39%) |
| **Antibiotic usage*** | |
| Main classes | |
| Cephalosporins | 22 (73%) |
| Carbapenems | 19 (63%) |
| Glycopeptides | 18 (60%) |
| Penicillins | 7 (23%) |
| Metrics | |
| LOT (median, range) | 15.5 (0–58) |
| DOT (median, range) | 22 (0–112) |

**Table S1: Clinical characteristics of study patients.** *Antibiotics usage percent values are calculated considering a total of 30 patients (see Materials and methods). HCT-CI, Hematopoietic Cell Transplantation-specific Comorbidity Index; DRI, Disease Risk Index; MRD, matched related donor; MUD, matched unrelated donor; MMUD, mismatched unrelated donor; LOT, length of therapy; DOT, days of therapy.

**Table S2**

| | Diversity stability | | | Composition stability | | |
|---|---|---|---|---|---|---|
| | **Coefficient** | **SE** | **P-value** | **Coefficient** | **SE** | **P-value** |
| **GCF** | | | | | | |
| Intercept | 0,9660 | 0,1007 | <0,0001 | 0,4468 | 0,1697 | 0,0146 |
| Cephalosporins | 0,0427 | 0,0969 | 0,6636 | -0,2937 | 0,1633 | 0,0848 |
| Carbapenems | 0,0088 | 0,0951 | 0,9269 | 0,0206 | 0,1603 | 0,8990 |
| Glycopeptides | -0,0895 | 0,1011 | 0,3848 | -0,1952 | 0,1704 | 0,2633 |
| Penicillins | 0,0872 | 0,1224 | 0,4830 | -0,3756 | 0,2063 | 0,0812 |
| DOT | -0,0057 | 0,0022 | **0,0172** | 0,0031 | 0,0037 | 0,4136 |
| **OM** | | | | | | |
| Intercept | 0,6794 | 0,1852 | 0,0014 | 0,0623 | 0,1978 | 0,7560 |
| Cephalosporins | 0,2011 | 0,1858 | 0,2913 | -0,0658 | 0,1985 | 0,7440 |
| Carbapenems | 0,1687 | 0,1706 | 0,3340 | -0,1948 | 0,1822 | 0,2970 |
| Glycopeptides | 0,2534 | 0,1877 | 0,1914 | 0,0171 | 0,2005 | 0,9330 |
| Penicillins | 0,1655 | 0,2158 | 0,4516 | 0,1565 | 0,2306 | 0,5050 |
| DOT | -0,0167 | 0,0046 | **0,0015** | 0,0019 | 0,0049 | 0,6970 |
| **SB** | | | | | | |
| Intercept | 0,7131 | 0,1829 | 0,0007 | 0,3928 | 0,1888 | 0,0488 |
| Cephalosporins | 0,2947 | 0,1764 | 0,1084 | -0,3677 | 0,1821 | 0,0553 |
| Carbapenems | 0,1024 | 0,1739 | 0,5617 | 0,1713 | 0,1795 | 0,3500 |
| Glycopeptides | -0,1365 | 0,1846 | 0,4673 | -0,4624 | 0,1906 | **0,0235** |
| Penicillins | 0,1890 | 0,2219 | 0,4031 | -0,1793 | 0,2290 | 0,4417 |
| DOT | -0,0085 | 0,0041 | **0,0467** | 0,0050 | 0,0042 | 0,2456 |

**Table S2: Multiple linear models testing prediction of oral microbiota stability by antibiotic usage.** Significant predictors are highlighted in bold. GCF, gingival crevicular fluid; OM, oral mucosa; SB, supragingival biofilm; DOT, days of therapy; SE, standard error.

**Table S3**

| | Outcome | N total | N group R | N group NR | % event R | % event NR | HR (95% CI) | P-value |
|---|---|---|---|---|---|---|---|---|
| **GCF recovery** | OS | 30 | 23 | 7 | 39 | 71 | 0,39 (0,13–1,16) | 0,0897 |
| | PFS | 28 | 22 | 6 | 36 | 50 | 0,51 (0,13–1,93) | 0,3197 |
| | Relapse | 28 | 22 | 6 | 36 | 50 | 0,66 (0,18–2,36) | 0,5200 |
| | TRD | 30 | 23 | 7 | 22 | 29 | 0,71 (0,14–3,66) | 0,6900 |
| **OM recovery** | OS | 29 | 20 | 9 | 25 | 89 | 0,17 (0,05–0,52) | **0,0020** |
| | PFS | 27 | 19 | 8 | 26 | 75 | 0,06 (0,01–0,34) | **0,0012** |
| | Relapse | 27 | 19 | 8 | 26 | 75 | 0,20 (0,06–0,69) | **0,0110** |
| | TRD | 29 | 20 | 9 | 10 | 44 | 0,19 (0,04–1,00) | 0,0500 |
| **SB recovery** | OS | 30 | 23 | 7 | 48 | 43 | 0,87 (0,24–3,14) | 0,8328 |
| | PFS | 28 | 22 | 6 | 41 | 33 | 1,15 (0,23–5,83) | 0,8622 |
| | Relapse | 28 | 22 | 6 | 41 | 33 | 1,54 (0,36–6,66) | 0,5600 |
| | TRD | 30 | 23 | 7 | 22 | 29 | 0,64 (0,12–3,50) | 0,6000 |

**Table S3: Univariate associations between oral microbiota recovery and clinical outcomes.** The total number of patients considered in each association is indicated (N total). The variation in N total per associations is caused by the unavailability of a sample essential for recovery evaluation or the exclusion of patients experiencing the event before recovery evaluation. Patients were grouped into recoverers (R) and non-recoverers (NR). The percentage of patients in each group experiencing the event is indicated. Q-value refers to the P-value adjusted for the number of oral sites tested. Significant associations are highlighted in bold. GCF, gingival crevicular fluid; OM, oral mucosa; SB, supragingival biofilm; OS, overall survival; PFS, progression-free survival; TRD, transplant-related death; HR, hazard ratio; CI, confidence interval.

**Table S4**

| Univariate associations with OS | | |
|---|---|---|
| | **HR (95% CI)** | **P-value** |
| Age in years | 1,01 (0,97–1,05) | 0,6223 |
| Sex (Female vs Male) | 1,28 (0,46–3,54) | 0,6399 |
| Underlying disease (AL vs Other) | 0,48 (0,17–1,32) | 0,1557 |
| HCT-CI (1-2 vs 0) | 1,67 (0,54–5,19) | 0,3746 |
| HCT-CI (≥3 vs 0) | 1,62 (0,40–6,49) | 0,4984 |
| DRI (H vs LI) | 3,93 (1,32–11,7) | **0,0139** |
| Conditioning intensity (M vs RI) | 0,27 (0,08–0,97) | **0,0443** |
| Total body irradiation (Yes vs No) | 1,76 (0,64–4,85) | 0,2779 |
| T-cell depletion (Yes vs No) | 2,79 (0,93–8,32) | 0,0659 |
| Graft source (BM vs PB) | 0,23 (0,05–1,02) | 0,0539 |
| Donor (HI vs MRD) | 0,89 (0,22–3,56) | 0,8675 |
| Donor (MUD/MMUD vs MRD) | 1,79 (0,50–6,37) | 0,3678 |
| Cephalosporins (Yes vs No) | 0,63 (0,21–1,88) | 0,4050 |
| Carbapenems (Yes vs No) | 2,46 (0,69–8,82) | 0,1674 |
| Glycopeptides (Yes vs No) | 1,36 (0,46–4,07) | 0,5803 |
| Penicillins (Yes vs No) | 1,49 (0,47–4,77) | 0,4984 |
| DOT | 1,04 (1,02–1,07) | **0,0006** |

**Table S4: Univariate associations between clinical parameters and overall survival (OS).** Significant associations are highlighted in bold. AL, acute leukemia; HCT-CI, Hematopoietic Cell Transplantation-specific Comorbidity Index; DRI, Disease Risk Index; H, high; LI, low-intermediate; M, myeloablative; RI, reduced intensity; BM, bone marrow; PB, peripheral blood; HI, haploidentical; MRD, matched related donor; MUD, matched unrelated donor; MMUD, mismatched unrelated donor; DOT, days of therapy; HR, hazard ratio; CI, confidence interval.

**Table S5**

| Univariate associations with PFS | | |
|---|---|---|
| | **HR (95% CI)** | **P-value** |
| Age in years | 0,99 (0,95–1,02) | 0,4348 |
| Sex (Female vs Male) | 2,41 (0,83–7,01) | 0,1079 |
| Underlying disease (AL vs Other) | 0,51 (0,18–1,50) | 0,2233 |
| HCT-CI (1-2 vs 0) | 1,57 (0,52–4,70) | 0,4219 |
| HCT-CI (≥3 vs 0) | 0,38 (0,05–3,08) | 0,3632 |
| DRI (H vs LI) | 7,12 (1,92–26,5) | **0,0034** |
| Conditioning intensity (M vs RI) | 0,65 (0,22–1,98) | 0,4533 |
| Total body irradiation (Yes vs No) | 2,03 (0,71–5,79) | 0,1877 |
| T-cell depletion (Yes vs No) | 2,75 (0,90–8,36) | 0,0745 |
| Graft source (BM vs PB) | 0,63 (0,20–2,01) | 0,4331 |
| Donor (HI vs MRD) | 2,03 (0,37–11,1) | 0,4166 |
| Donor (MUD/MMUD vs MRD) | 4,03 (0,85–19,1) | 0,0792 |
| Cephalosporins (Yes vs No) | 0,50 (0,16–1,53) | 0,2230 |
| Carbapenems (Yes vs No) | 1,50 (0,46–4,94) | 0,5023 |
| Glycopeptides (Yes vs No) | 1,68 (0,52–5,46) | 0,3892 |
| Penicillins (Yes vs No) | 0,65 (0,14–2,94) | 0,5779 |
| DOT | 1,01 (0,98–1,05) | 0,3782 |

**Table S5: Univariate associations between clinical parameters and progression-free survival (PFS).** Significant associations are highlighted in bold. AL, acute leukemia; HCT-CI, Hematopoietic Cell Transplantation-specific Comorbidity Index; DRI, Disease Risk Index; H, high; LI, low-intermediate; M, myeloablative; RI, reduced intensity; BM, bone marrow; PB, peripheral blood; HI, haploidentical; MRD, matched related donor; MUD, matched unrelated donor; MMUD, mismatched unrelated donor; DOT, days of therapy; HR, hazard ratio; CI, confidence interval.

**Table S6**

| Univariate associations with relapse | | |
|---|---|---|
| | **HR (95% CI)** | **P-value** |
| Age in years | 0,98 (0,95–1,02) | 0,3000 |
| Sex (Female vs Male) | 2,48 (0,89–6,85) | 0,0810 |
| Underlying disease (AL vs Other) | 0,61 (0,22–1,68) | 0,3400 |
| HCT-CI (1-2 vs 0) | 1,77 (0,61–5,18) | 0,2900 |
| HCT-CI (≥3 vs 0) | 0,31 (0,04–2,40) | 0,2600 |
| DRI (H vs LI) | 5,96 (1,94–18,3) | **0,0018** |
| Conditioning intensity (M vs RI) | 0,79 (0,28–2,20) | 0,6500 |
| Total body irradiation (Yes vs No) | 2,09 (0,77–5,68) | 0,1500 |
| T-cell depletion (Yes vs No) | 2,20 (0,79–6,14) | 0,1300 |
| Graft source (BM vs PB) | 0,79 (0,26–2,39) | 0,6800 |
| Donor (HI vs MRD) | 2,07 (0,39–11,1) | 0,4000 |
| Donor (MUD/MMUD vs MRD) | 3,84 (0,83–17,7) | 0,0840 |
| Cephalosporins (Yes vs No) | 0,52 (0,19–1,41) | 0,2000 |
| Carbapenems (Yes vs No) | 1,16 (0,34–3,93) | 0,8200 |
| Glycopeptides (Yes vs No) | 1,48 (0,46–4,72) | 0,5100 |
| Penicillins (Yes vs No) | 0,47 (0,12–1,76) | 0,2600 |
| DOT | 0,99 (0,98–1,01) | 0,5200 |

**Table S6: Univariate associations between clinical parameters and risk of relapse.** Significant associations are highlighted in bold. AL, acute leukemia; HCT-CI, Hematopoietic Cell Transplantation-specific Comorbidity Index; DRI, Disease Risk Index; H, high; LI, low-intermediate; M, myeloablative; RI, reduced intensity; BM, bone marrow; PB, peripheral blood; HI, haploidentical; MRD, matched related donor; MUD, matched unrelated donor; MMUD, mismatched unrelated donor; DOT, days of therapy; HR, hazard ratio; CI, confidence interval.

**Table S7**

| Univariate associations with TRD | | |
|---|---|---|
| | **HR (95% CI)** | **P-value** |
| Age in years | 1,00 (0,95–1,05) | 0,9600 |
| Sex (Female vs Male) | 0,51 (0,10–2,5) | 0,4000 |
| Underlying disease (AL vs Other) | 0,49 (0,11–2,07) | 0,3300 |
| HCT-CI (1-2 vs 0) | 2,45 (0,46–13,2) | 0,3000 |
| HCT-CI (≥3 vs 0) | 3,36 (0,52–21,8) | 0,2000 |
| DRI (H vs LI) | 1,00 (0,24–4,25) | 1 |
| Conditioning intensity (M vs RI) | 0,63 (0,12–3,16) | 0,5700 |
| Total body irradiation (Yes vs No) | 1,30 (0,32–5,39) | 0,7100 |
| T-cell depletion (Yes vs No) | 1,40 (0,34–5,73) | 0,6400 |
| Graft source (BM vs PB) | 0,32 (0,04–2,42) | 0,2700 |
| Donor (HI vs MRD) | 0,97 (0,15–6,40) | 0,9700 |
| Donor (MUD/MMUD vs MRD) | 1,43 (0,29–6,94) | 0,6600 |
| Cephalosporins (Yes vs No) | 0,89 (0,19–4,22) | 0,8800 |
| Carbapenems (Yes vs No) | 3,74 (0,47–29,6) | 0,2100 |
| Glycopeptides (Yes vs No) | 1,88 (0,41–8,68) | 0,4200 |
| Penicillins (Yes vs No) | 3,06 (0,69–13,6) | 0,1400 |
| DOT | 1,06 (1,03–1,08) | **<0.0001** |

**Table S7: Univariate associations between clinical parameters and risk of transplant-related death (TRD).** Significant associations are highlighted in bold. AL, acute leukemia; HCT-CI, Hematopoietic Cell Transplantation-specific Comorbidity Index; DRI, Disease Risk Index; H, high; LI, low-intermediate; M, myeloablative; RI, reduced intensity; BM, bone marrow; PB, peripheral blood; HI, haploidentical; MRD, matched related donor; MUD, matched unrelated donor; MMUD, mismatched unrelated donor; DOT, days of therapy; HR, hazard ratio; CI, confidence interval.

**Table S8**

| Multivariate associations | | |
|---|---|---|
| | **HR (95% CI)** | **P-value** |
| **OS** | | |
| OM recovery (R vs NR) | 0,09 (0,02–0,35) | **0,0006** |
| DRI (H vs LI) | 6,34 (1,58–25,5) | **0,0092** |
| Conditioning intensity (M vs RI) | 0,16 (0,03–0,88) | **0,0353** |
| DOT | 1,04 (1,00–1,09) | **0,0457** |
| **PFS** | | |
| OM recovery (R vs NR) | 0,09 (0,02–0,49) | **0,0052** |
| DRI (H vs LI) | 3,61 (0,85–15,2) | 0,0807 |
| **Relapse** | | |
| OM recovery (R vs NR) | 0,19 (0,06–0,55) | **0,0025** |
| DRI (H vs LI) | 4,69 (1,47–15,0) | **0,0090** |

**Table S8: Multivariate associations of oral microbiota recovery and clinical parameters with clinical outcomes.** Clinical parameters significantly associated with the outcome in the univariate models (Table S4–6) were used to adjust the significant univariate associations between oral microbiota recovery and clinical outcomes (Table S3). Significant associations are highlighted in bold. OM, oral mucosa; R, recoverers; NR, non-recoverers; DRI, Disease Risk Index; H, high; LI, low-intermediate; M, myeloablative; RI, reduced intensity; DOT, days of therapy; OS, overall survival; PFS, progression-free survival; HR, hazard ratio; CI, confidence interval.

**Table S9**

| | N or median (IQR) | | P-value |
|---|---|---|---|
| | R | NR | |
| **Age in years** | 51,1 (15,1) | 51,1 (21,5) | 0,6268 |
| **Sex** | | | 0,6942 |
| Female | 9 | 3 | |
| Male | 11 | 6 | |
| **Underlying disease** | | | 0,4223 |
| Acute leukemia | 13 | 4 | |
| Other | 7 | 5 | |
| **HCT-CI** | | | 0,4758 |
| 0 | 12 | 3 | |
| 1-2 | 5 | 4 | |
| ≥3 | 3 | 2 | |
| **DRI** | | | 1 |
| Low-intermediate | 12 | 5 | |
| High | 8 | 4 | |
| **Conditioning intensity** | | | 0,6942 |
| Reduced intensity | 11 | 6 | |
| Myeloablative | 9 | 3 | |
| **Total body irradiation** | | | 0,3962 |
| Yes | 5 | 4 | |
| No | 15 | 5 | |
| **T-cell depletion** | | | 0,427 |
| Yes | 9 | 6 | |
| No | 11 | 3 | |
| **Graft source** | | | 0,4311 |
| Bone marrow | 8 | 2 | |
| Peripheral blood | 12 | 7 | |
| **Donor** | | | 0,535 |
| MRD | 7 | 2 | |
| Haploidentical | 7 | 2 | |
| MUD/MMUD | 6 | 5 | |
| **Cephalosporins** | | | 0,2089 |
| Yes | 16 | 5 | |
| No | 4 | 4 | |

| | | | |
|---|---|---|---|
| **Carbapenems** | | | 0,4118 |
| Yes | 11 | 7 | |
| No | 9 | 2 | |
| **Glycopeptides** | | | 0,6942 |
| Yes | 11 | 6 | |
| No | 9 | 3 | |
| **Penicillins** | | | 1 |
| Yes | 4 | 2 | |
| No | 16 | 7 | |
| **LOT** | 14,5 (8,75) | 19 (12) | 0,2986 |
| **DOT** | 21 (19,25) | 22 (23) | 0,4639 |

**Table S9: Associations between clinical parameters and oral mucosa (OM) microbiota recovery.** The Fisher's exact test and the Mann-Whitney U test were used for categorical and continuous variables, respectively. For categorical variables, the contingency table is shown. For numerical variables, the median value and the interquartile range (IQR) for each group are shown. R, OM recoverers; NR, OM non-recoverers; HCT-CI, Hematopoietic Cell Transplantation-specific Comorbidity Index; DRI, Disease Risk Index; MRD, matched related donor; MUD, matched unrelated donor; MMUD, mismatched unrelated donor; LOT, length of therapy; DOT, days of therapy.

#### SUPPLEMENTARY FIGURES

##### Figure S1

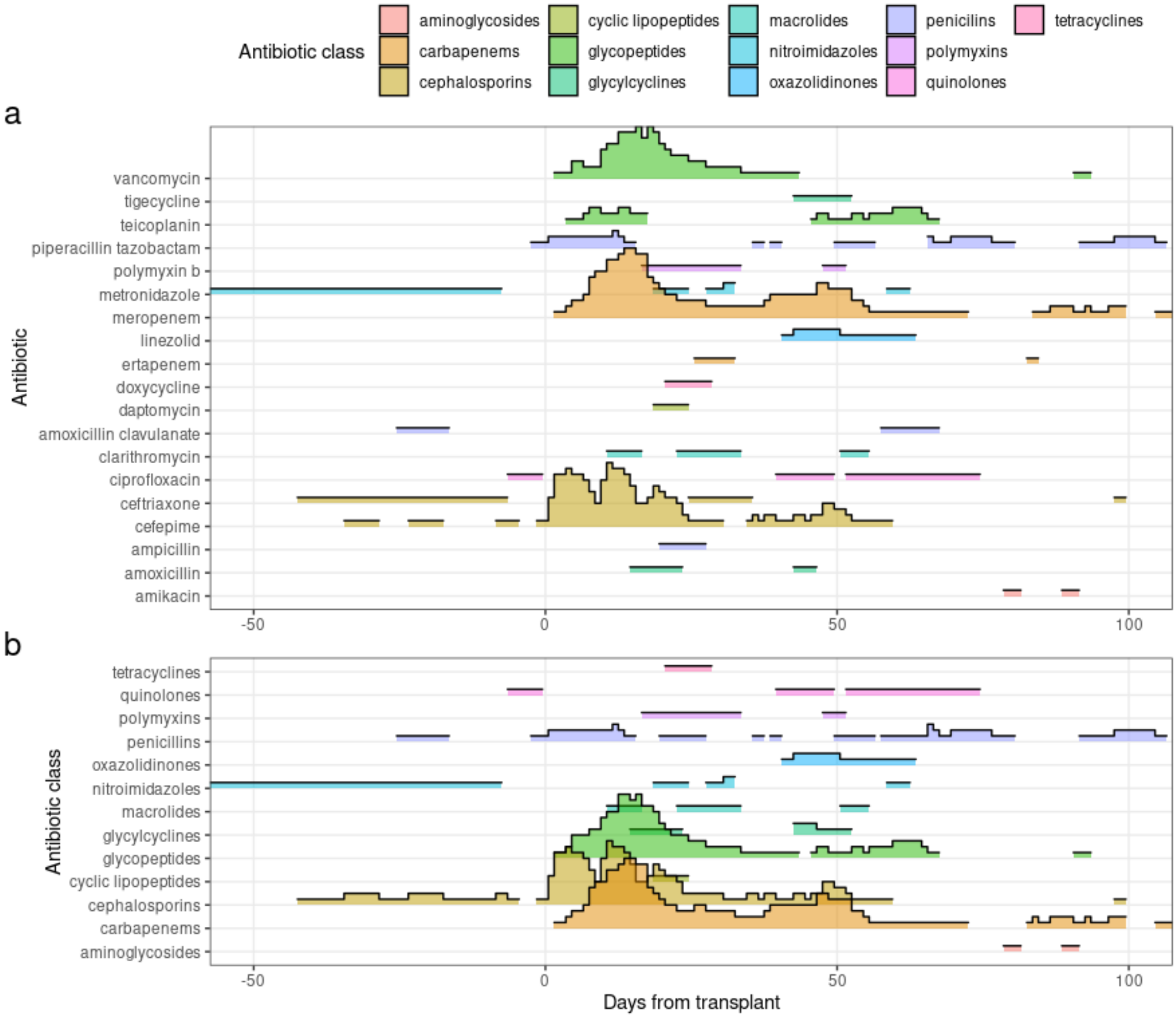


**Figure S1: a** Ridgeline plot of the antibiotic agents used by the cohort in relation to stem-cell infusion. **b** Ridgeline plot of the antibiotic classes used by the cohort in relation to stem-cell infusion.

**Figure S2**

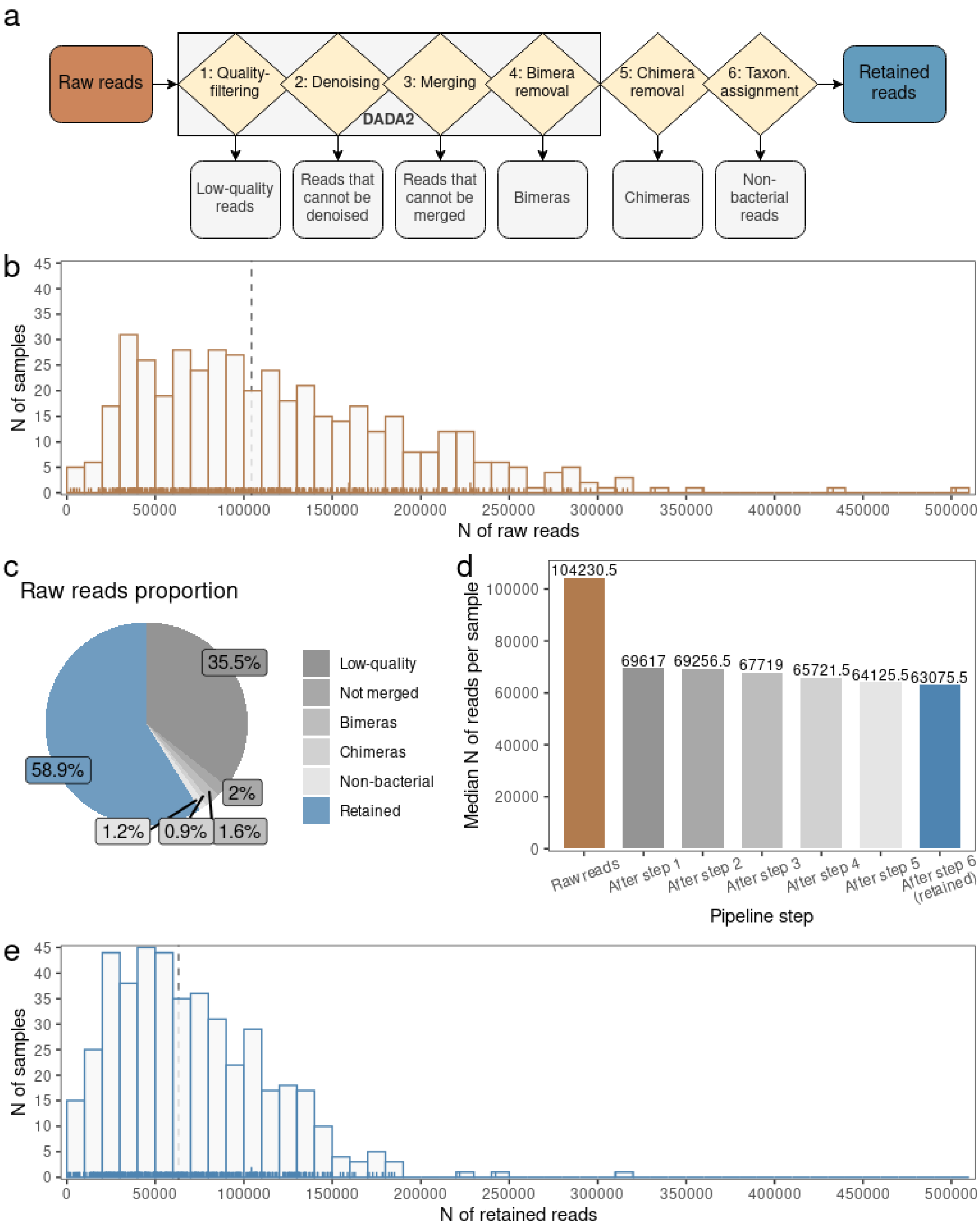


**Figure S2:** **a** Read processing pipeline scheme. **b** Histogram with the number of raw reads per sample. **c** Proportion of reads discarded at each pipeline step. **d** Median number of reads per sample at each pipeline step. **e** Histogram with the number of retained reads after running the read processing pipeline.

**Figure S3**

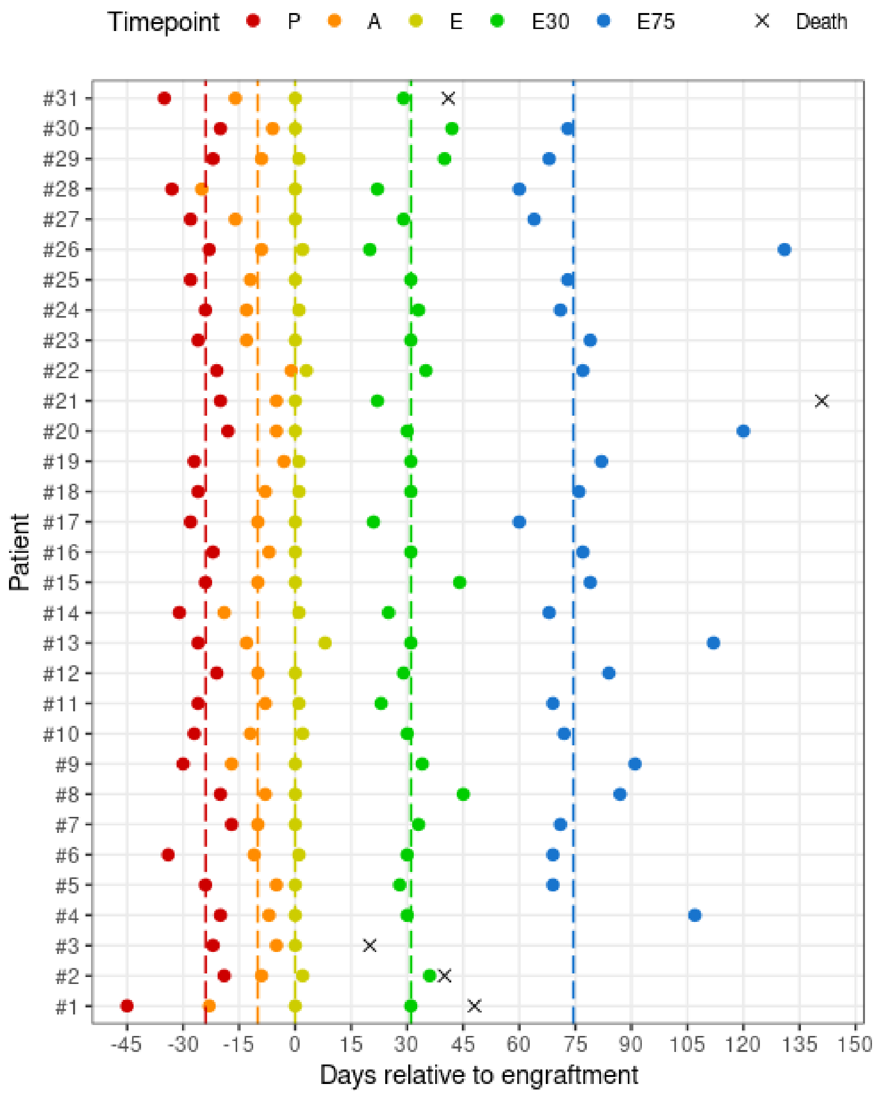


**Figure S3:** Sampling times for each patient in relation to engraftment day. Vertical dashed lines indicate the median sampling time per timepoint. P, preconditioning; A, aplasia; E, engraftment; E30, 30 days after engraftment; E75, 75 days after engraftment.

**Figure S4**

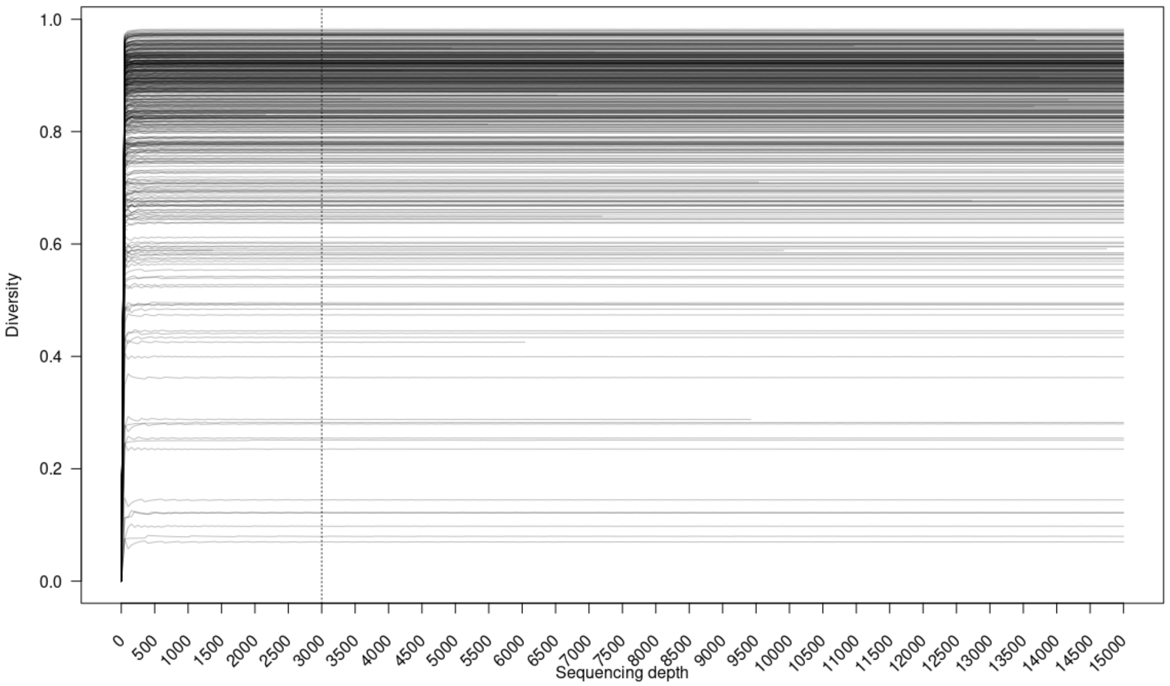


**Figure S4:** Gini-Simpson rarefaction curves per sample. Reads were selected by scaling with ranked subsampling (SRS) at incremental steps of 50 reads. The plot was limited to 15,000 reads.

#### Figure S5

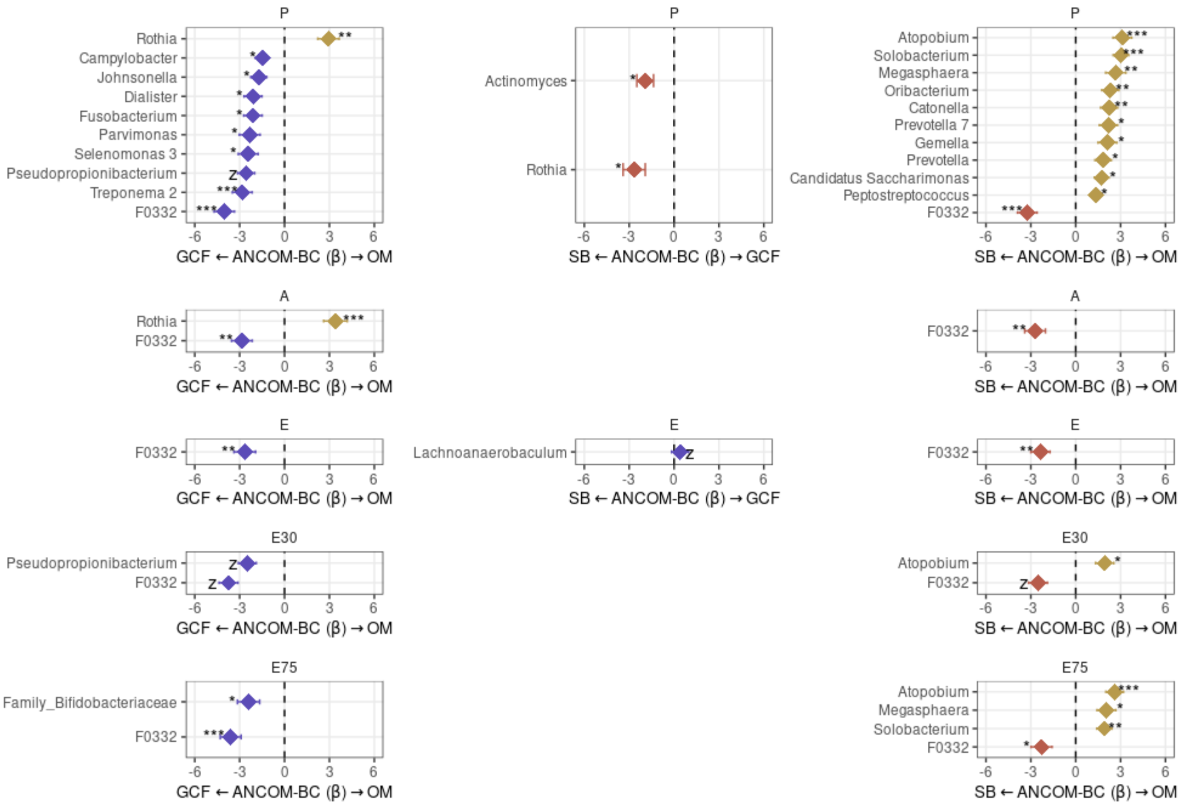


**Figure S5:** Differentially abundant genera (ANCOM-BC) between oral sites at each timepoint. GCF, gingival crevicular fluid; OM, oral mucosa; SB, supragingival biofilm; P, preconditioning; A, aplasia; E, engraftment; E30, 30 days after engraftment; E75, 75 days after engraftment; *, q-value < 0.05; **, q-value < 0.01; ***, q-value < 0.001; z, ANCOM-BC structural zero.

.

#### Figure S6

a

Resistance (P → E)

$$RS = 1 - \frac{2|\alpha_p - \alpha_e|}{\alpha_p + |\alpha_p - \alpha_e|}$$

Resilience (E → E30)

$$RL = \left(\frac{2|\alpha_p - \alpha_e|}{|\alpha_p - \alpha_e| + |\alpha_p - \alpha_{e30}|} - 1\right) \div t$$

Stability (P → E30)

$$S = 1 - \frac{4|\alpha_p - \alpha_e||\alpha_p - \alpha_{e30}|}{(\alpha_p + |\alpha_p - \alpha_e|)(|\alpha_p - \alpha_e| + |\alpha_p - \alpha_{e30}|)}$$

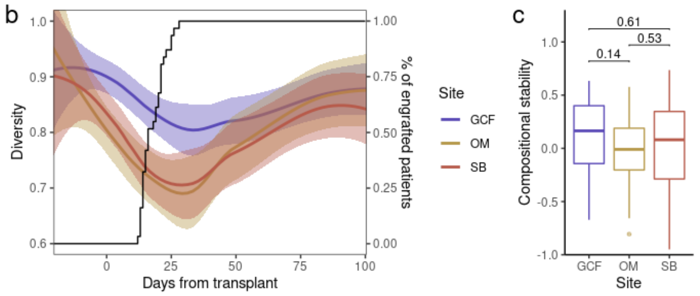


**Figure S6: a** Diversity resistance, resilience, and stability expressions (see Methods). **b** Smoothed trend-line of diversity (Gini-Simpson) in each oral site (left y-axis) and percentage of engrafted patients (right y-axis) per day from stem-cell infusion. Shaded areas represent 95% confidence intervals. **c** Compositional stability (see Methods) per oral site. Mann-Whitney U test was used. GCF, gingival crevicular fluid; OM, oral mucosa; SB, supragingival biofilm.

### Figure S7

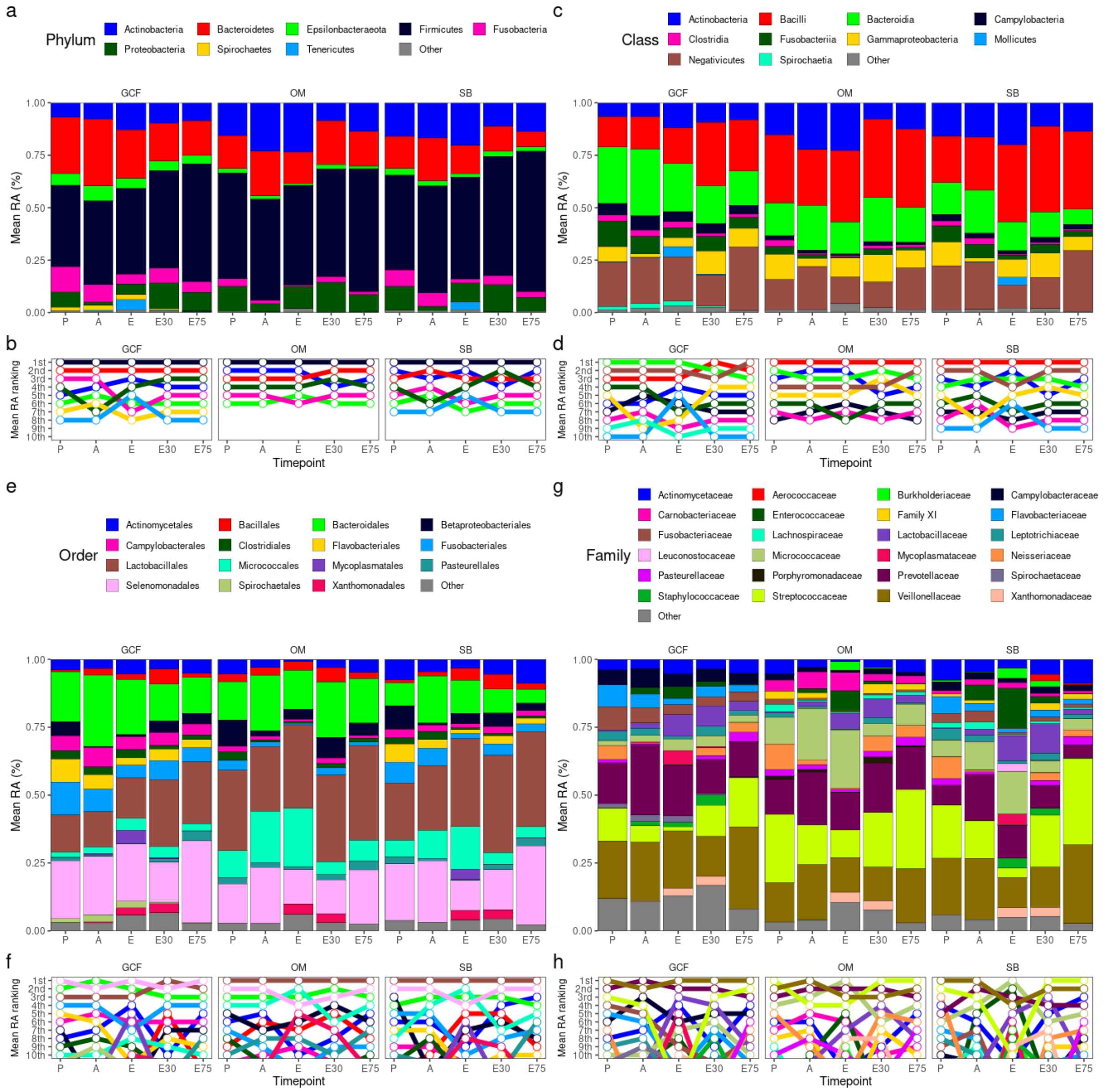


**Figure S7: a, c, e, g** Mean phyla (a), class (c), order (e), and family (g) relative abundances (RA) per timepoint for each oral site. Taxa with >2% mean RA in any combination of oral site and timepoint are shown. **b, d, f, h** Mean phyla (b), class (d), order (f), and family (g) RA ranking per timepoint for each oral site. Top-10 taxa are shown. GCF, gingival crevicular fluid; OM, oral mucosa; SB, supragingival biofilm; P, preconditioning; A, aplasia; E, engraftment; E30, 30 days after engraftment; E75, 75 days after engraftment.

**Figure S8**

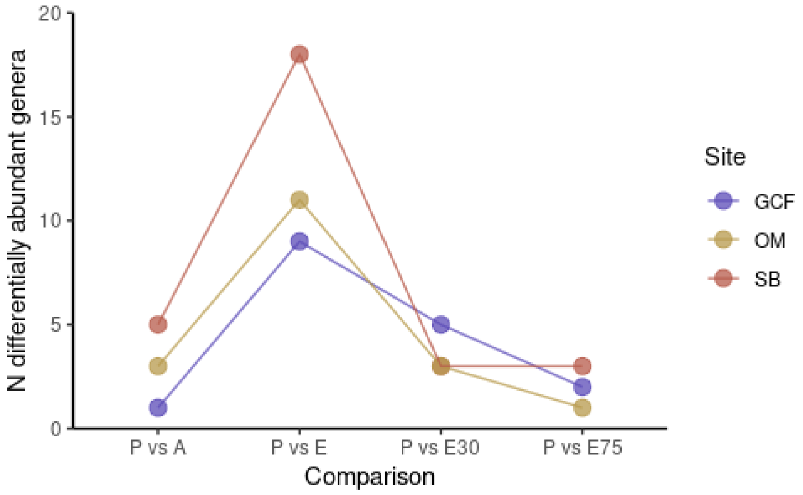


**Figure S8:** Number of differentially abundant genera (ANCOM-BC) between preconditioning (P) and other timepoints for each site. GCF, gingival crevicular fluid; OM, oral mucosa; SB, supragingival biofilm; A, aplasia; E, engraftment; E30, 30 days after engraftment; E75, 75 days after engraftment.

**Figure S9**

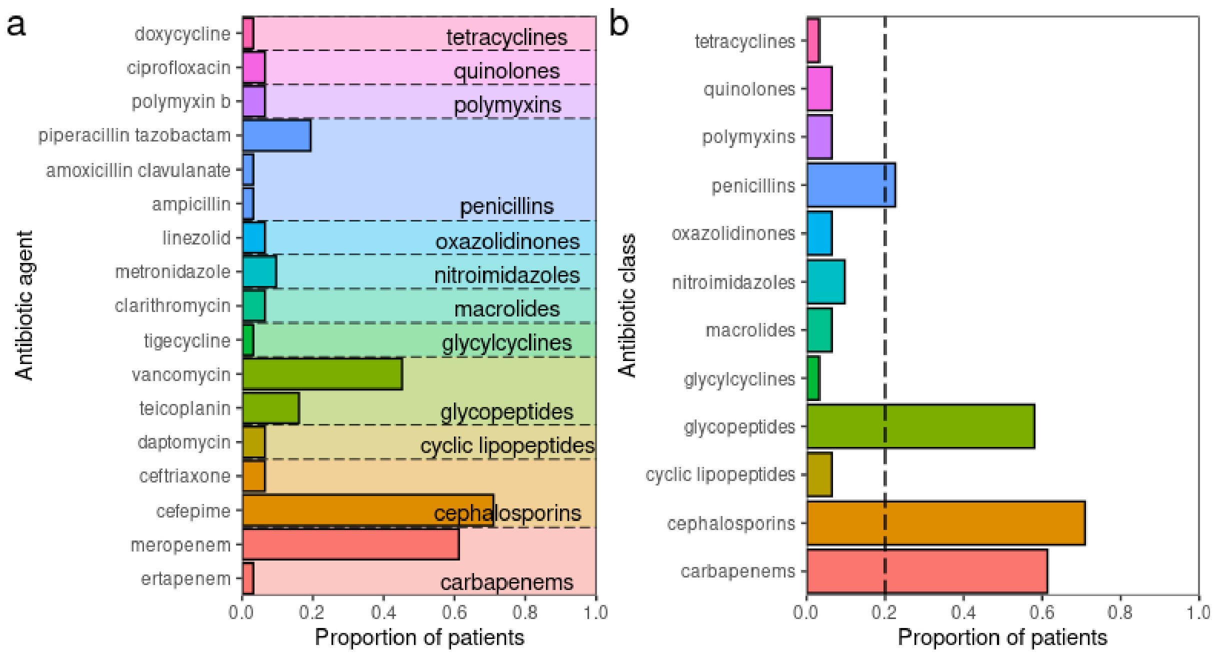


**Figure S9: a** Proportion of patients using each antibiotic agent between preconditioning (P) and 30 days after engraftment (E30). Respective antibiotic classes are indicated. **b** Proportion of patients using each antibiotic class between P and E30. Vertical dashed line indicates the proportion of 20%.

#### Figure S10

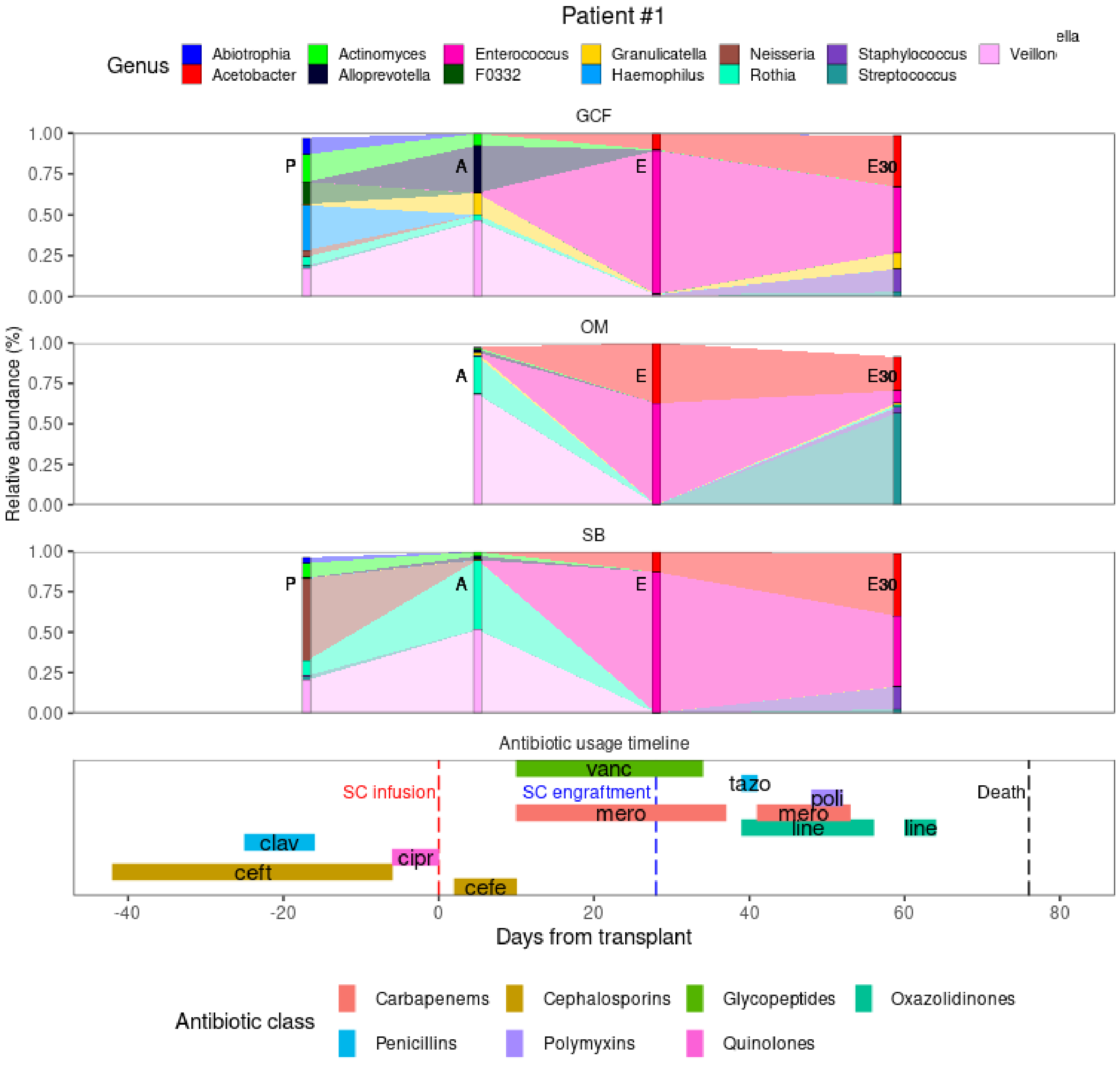


**Figure S10:** Patient #1: genera relative abundance dynamics for each oral site (top) and antibiotic usage timeline (bottom). Genera with >1% mean relative abundance in any combination of oral site and timepoint are shown. GCF, gingival crevicular fluid; OM, oral mucosa; SB, supragingival biofilm; A, aplasia; E, engraftment; E75, 75 days after engraftment; SC, stem-cell; vanc, vancomycin; tazo, piperacillin tazobactam; poli, polymyxin B; mero, meropenem; line, linezolid; clav, amoxicillin clavulanate; cipr, ciprofloxacin; ceft, ceftriaxone; cefe, cefepime.

.

**Figure S11**

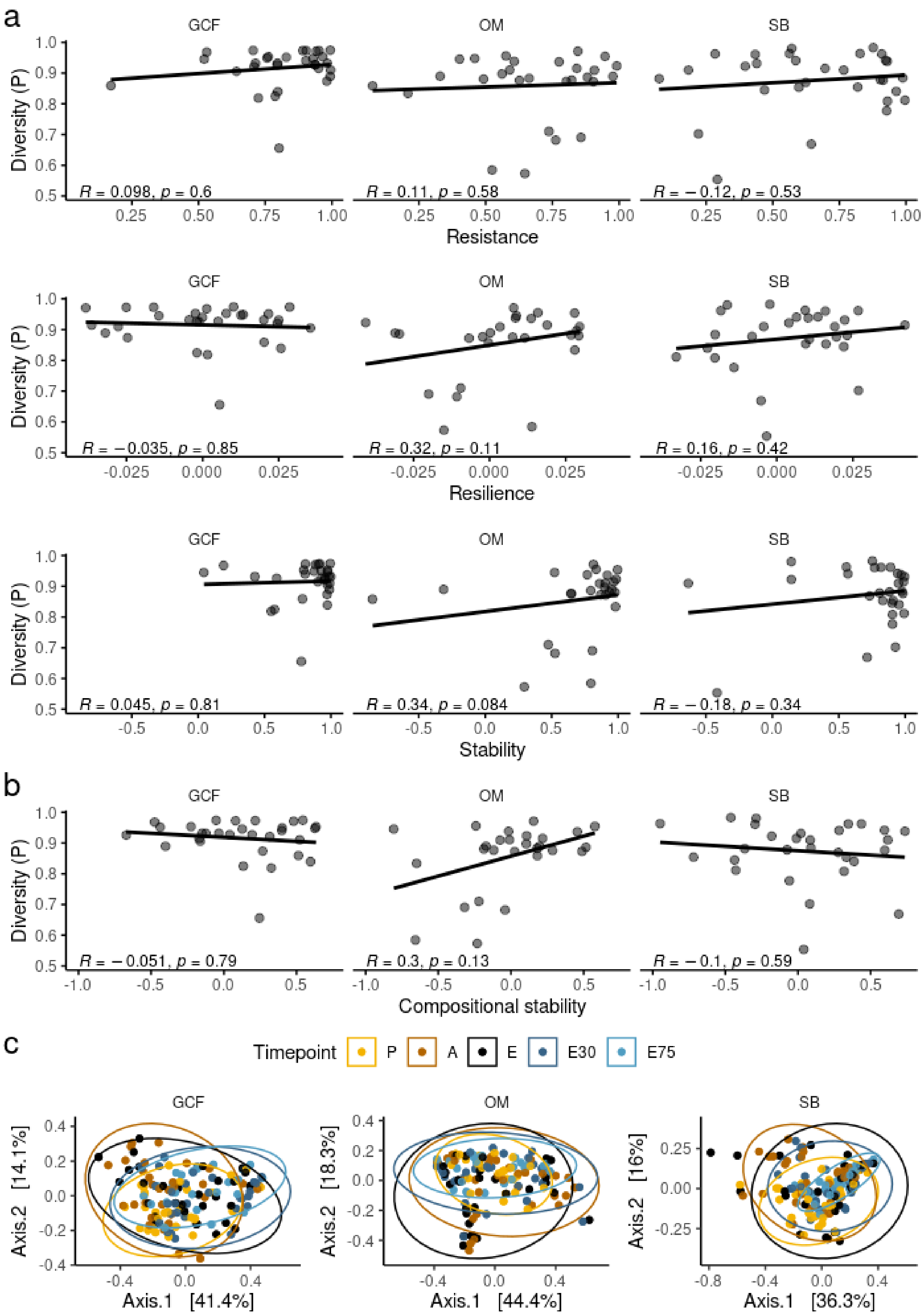
a
GCF
OM
SB
Diversity (P)
R = 0.098, p = 0.6
R = 0.11, p = 0.58
R = −0.12, p = 0.53
Resistance
R = −0.035, p = 0.85
R = 0.32, p = 0.11
R = 0.16, p = 0.42
Resilience
R = 0.045, p = 0.81
R = 0.34, p = 0.084
R = −0.18, p = 0.34
Stability
b
R = −0.051, p = 0.79
R = 0.3, p = 0.13
R = −0.1, p = 0.59
Compositional stability
c
Timepoint
P
A
E
E30
E75
Axis.2 [14.1%]
Axis.1 [41.4%]
Axis.2 [18.3%]
Axis.1 [44.4%]
Axis.2 [16%]
Axis.1 [36.3%]

**Figure S11: a** Correlation between diversity (Gini-Simpson) at preconditioning (P) and diversity resistance, resilience, or stability for each oral site. Spearman's rank correlation test was used. **b** Correlation between diversity (Gini-Simpson) at P and compositional stability for each oral site. Spearman's rank correlation test was used. **c** Principal Coordinate Analysis (PCoA) of microbiota distances (weighted UniFrac) between timepoints for each oral site. Ellipsoids indicate 95% confidence intervals. GCF, gingival crevicular fluid; OM, oral mucosa; SB, supragingival biofilm; P, preconditioning; A, aplasia; E, engraftment; E30, 30 days after engraftment; E75, 75 days after engraftment.

**Figure S12**

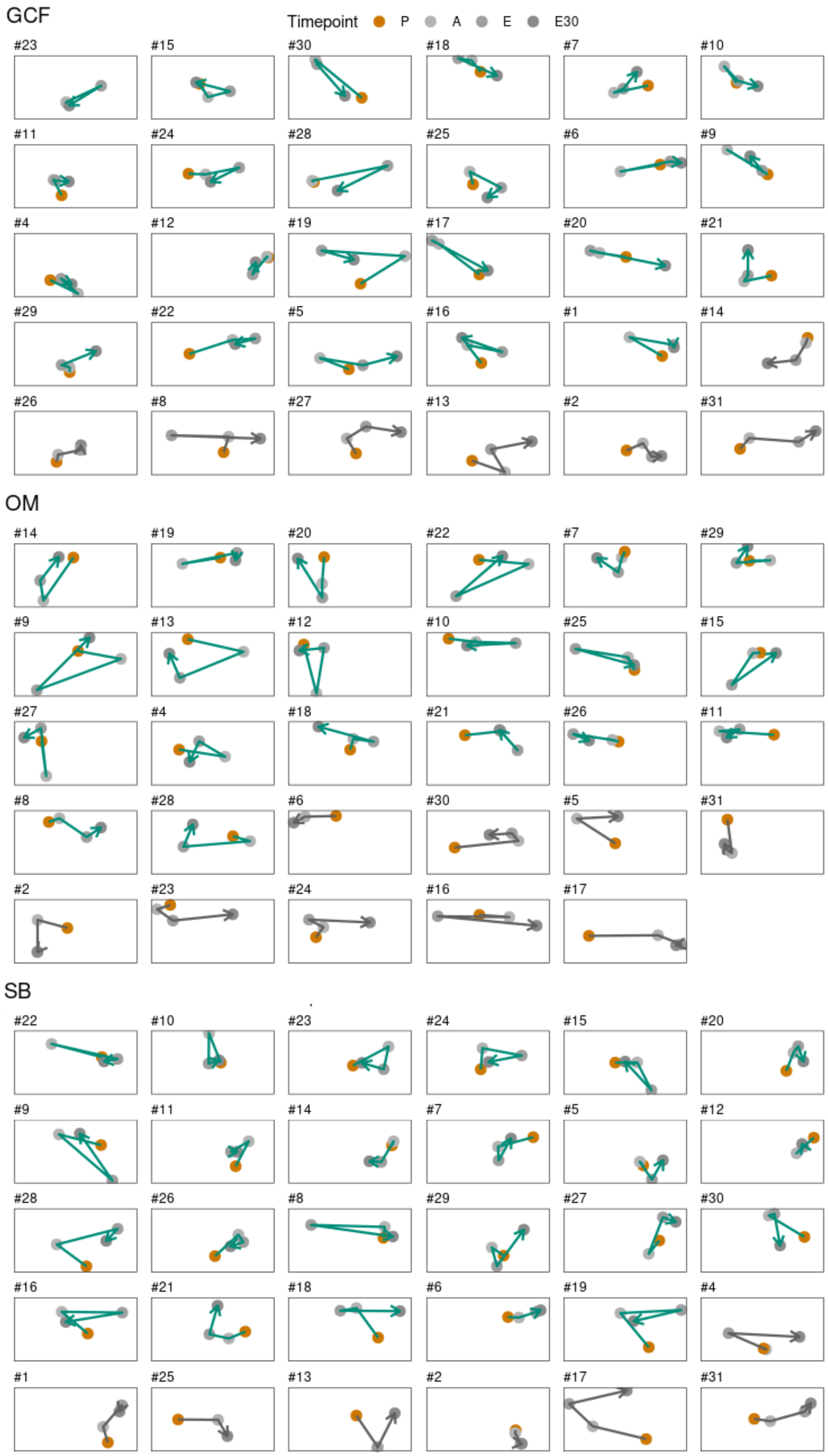

GCF
Timepoint P A E E30
#23
#15
#30
#18
#7
#10
#11
#24
#28
#25
#6
#9
#4
#12
#19
#17
#20
#21
#29
#22
#5
#16
#1
#14
#26
#8
#27
#13
#2
#31
OM
#14
#19
#20
#22
#7
#29
#9
#13
#12
#10
#25
#15
#27
#4
#18
#21
#26
#11
#8
#28
#6
#30
#5
#31
#2
#23
#24
#16
#17
SB
#22
#10
#23
#24
#15
#20
#9
#11
#14
#7
#5
#12
#28
#26
#8
#29
#27
#30
#16
#21
#18
#6
#19
#4
#1
#25
#13
#2
#17
#31

**Figure S12:** Principal Coordinate Analysis (PCoA) with microbiota trajectories for each patient in each oral site. Recovery trajectories are shown in teal and non-recovery in grey. GCF, gingival crevicular fluid; OM, oral mucosa; SB, supragingival biofilm; P, preconditioning; A, aplasia; E, engraftment; E30, 30 days after engraftment; E75, 75 days after engraftment.

#### Figure S13

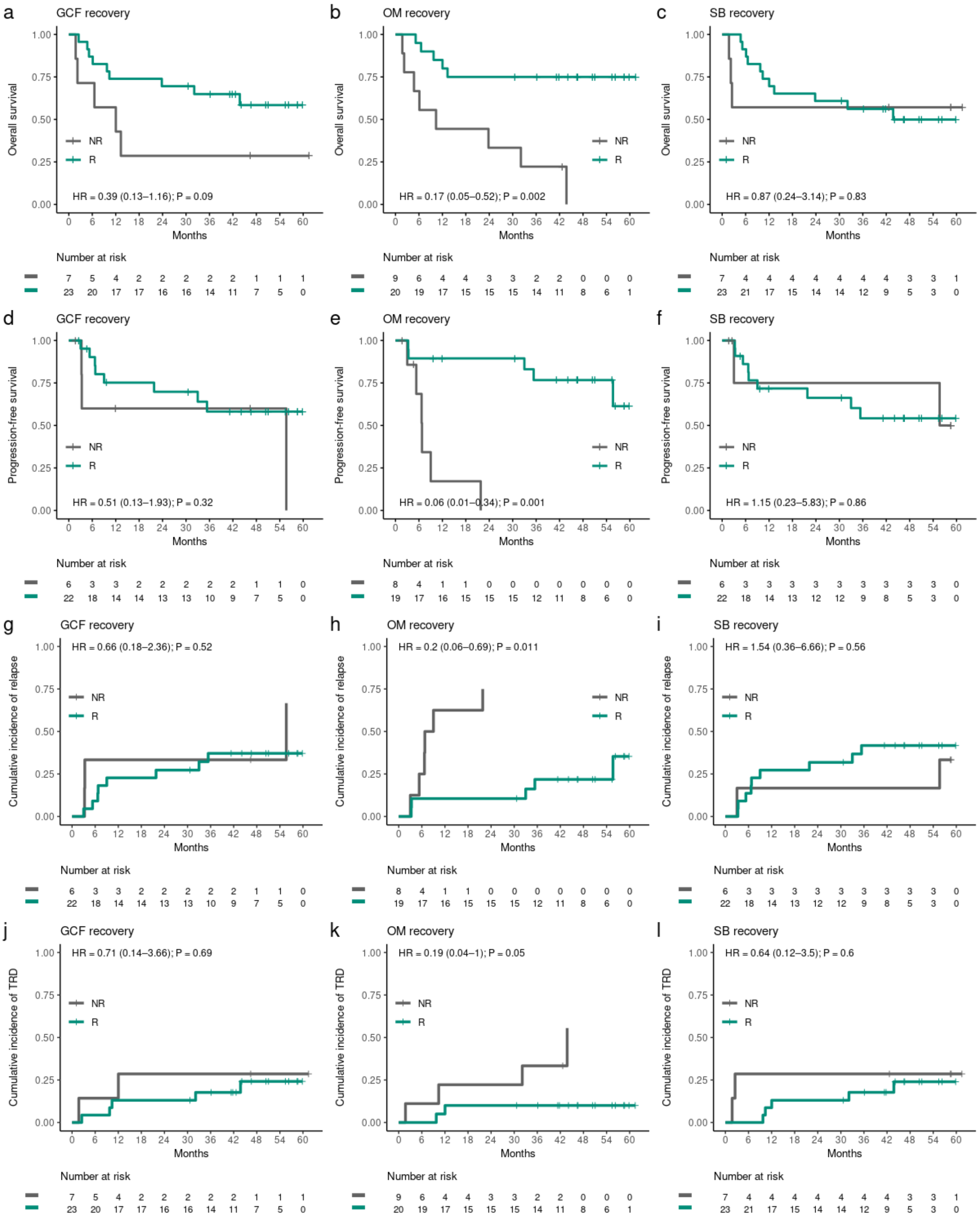


**Figure S13: a-c** Kaplan-Meier curves comparing overall survival among recoverers (R) and non-recoverers (NR) for each oral site. **d-f** Kaplan-Meier curves comparing progression-free survival among R and NR for each oral site. **g-i** Cumulative incidence curves of relapse among R and NR for each oral site. **j-l** Cumulative incidence curves of transplant-related death (TRD) among R and NR for each oral site. GCF, gingival crevicular fluid; OM, oral mucosa; SB, supragingival biofilm. HR, hazard ratio.

**Figure S14**

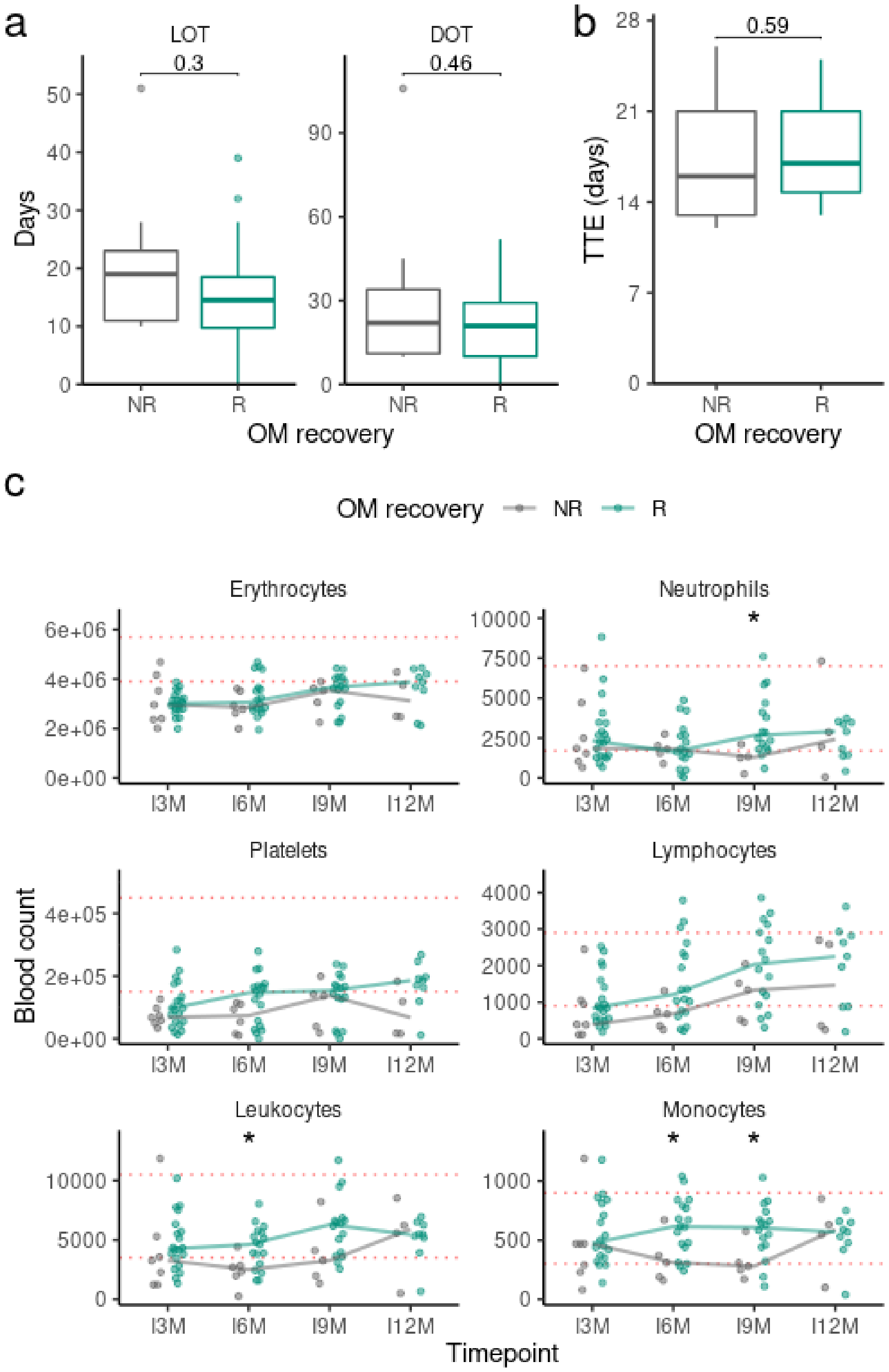


**Figure S14: a** Time of antibiotic administration (LOT: length of therapy; DOT: days of therapy) among oral mucosa (OM) recoverers (R) and non-recoverers (NR). **b** Time to engraftment (TTE) in days among OM R and NR. **c** Blood cell counts among OM R and NR per timepoint (described below) for each blood cell type. Red dotted horizontal lines indicate normal counts (within reference values). Solid lines indicate median values at each timepoint. Mann-Whitney U test was used. I3M, 3 months after stem-cell infusion; I6M, 6 months after stem-cell infusion; I9M, 9 months after stem-cell infusion; I12M, 12 months after stem-cell infusion; *, P-value < 0.05.
