## Additional file 3: Supplementary methods for "Longitudinal analysis at three oral sites links oral microbiota to clinical outcomes in allogeneic hematopoietic stem-cell transplant"

### Diversity resistance, resilience, and stability

The resistance ($RS$) of a microbiota parameter $y$ measures the level of alteration undergone by $y$ during a perturbation $l$. As proposed by Orwin & Wardle (2004), let $y_0$ be $y$ at baseline and $y_l$ be $y$ immediately after $l$, $y$ resistance to $l$ can be measured by:

$$RS = 1 - \frac{2|y_0 - y_l|}{y_0 + |y_0 - y_l|} \quad (1)$$

Rewriting this expression in the context of this study, with $y_0$ being the diversity at preconditioning ($\alpha_p$) and $y_l$ the diversity at engraftment ($\alpha_e$), diversity resistance to allo-HSCT can be calculated for each patient as follows:

$$RS = 1 - \frac{2|\alpha_p - \alpha_e|}{\alpha_p + |\alpha_p - \alpha_e|} \quad (2)$$

Orwin & Wardle further proposed an expression for $y$ resilience ($RL$), which refers to the rate of change of $y$ towards $y_0$ after $l$. Let $y_f$ be $y$ after a period of time $t$ after $l$ (i.e., $t = t_f - t_l$), $y$ resilience at $t$ can be measured by:

$$RL = \left(\frac{2|y_0 - y_l|}{|y_0 - y_l| + |y_0 - y_f|} - 1\right) \div t \quad (3)$$

Rewriting this expression in the context of this study, with $y_f$ being the diversity at 30 days after engraftment ($\alpha_{e30}$) and $t$ being the interval in days between the engraftment and the 30 days after engraftment sampling times ($t = t_{e30} - t_e$), diversity resilience to allo-HSCT at 30 days after engraftment can be calculated for each patient as follows:

$$RL = \left(\frac{2|\alpha_p - \alpha_e|}{|\alpha_p - \alpha_e| + |\alpha_p - \alpha_{e30}|} - 1\right) \div t \quad (4)$$

Orwin & Wardle did not propose an expression for $y$ stability ($S$). However, because stability is by definition composed of resistance and resilience (Shade et al., 2012), it follows that $S \propto RS + RL$, so that an expression for $y$ stability can be algebraically derived.

First, it must be noted that $RS$ is a unitless quantity, while $RL$ is a rate ($time^{-1}$). Therefore, in order to combine $RS$ and $RL$, $RL$ must be multiplied by a factor $b \propto t$ in the new stability expression.

$$S = RS + RL \times b \quad (5)$$

In order to find $b$, let us expand (5) using (2) and (4):

$$S = 1 - \frac{2|y_0 - y_l|}{y_0 + |y_0 - y_l|} + \left(\frac{2|y_0 - y_l|}{|y_0 - y_l| + |y_0 - y_f|} - 1\right) \times \frac{b}{t} \quad (6)$$

Now, it is reasonable to impose that when $y$ recovers to baseline levels after $t$, $y$ stability is maximum: $y_f = y_0 \Rightarrow S = 1$. Using this information in (6), it is possible to solve the equation for $b$:

$$1 = 1 - \frac{2|y_0 - y_l|}{y_0 + |y_0 - y_l|} + \left(\frac{2|y_0 - y_l|}{|y_0 - y_l| + |y_0 - y_0|} - 1\right) \times \frac{b}{t} \quad (7)$$

Resulting that

$$b = \frac{2t|y_0 - y_l|}{y_0 + |y_0 - y_l|} \quad (8)$$

As desired, $b \propto t$. Replacing $b$ in (6), it results that $y$ stability is given by:

$$S = 1 - \frac{4|y_0 - y_l||y_0 - y_f|}{(y_0 + |y_0 - y_l|)(|y_0 - y_l| + |y_0 - y_f|)} \quad (9)$$

Which in the context of this study can be rewritten as follows:

$$S = 1 - \frac{4|\alpha_p - \alpha_e||\alpha_p - \alpha_{e30}|}{(\alpha_p + |\alpha_p - \alpha_e|)(|\alpha_p - \alpha_e| + |\alpha_p - \alpha_{e30}|)} \quad (10)$$

**Compositional stability**

In analogy to diversity stability, which is calculated based on $\alpha_p$, $\alpha_e$, and $\alpha_{e30}$, compositional stability is evaluated by considering preconditioning, engraftment, and 30 days after engraftment samples. Specifically, let $C$ be the area in the compositional space enclosed by the convex hull of these samples. Compositional stability is calculated as $1 - C$.

**Multiple linear regression**

Multiple linear regression was used to evaluate whether antibiotic usage parameters predicted diversity and compositional stability (S). Separate models for each site were run with the *lm* function from the *stat* R package. Due to the high collinearity between DOT and LOT, only DOT and the main antibiotic classes were included as predictors in the models: $S \sim cephalosporins + carbapenems + glycopeptides + penicillins + DOT$.

**Taxonomic nomenclature homogenization**

Either due to a lack of taxonomic resolution or incomplete annotated taxonomic information in the 16S rRNA database used, assigned taxonomies may contain generic proxies at low taxonomic ranks, such as “uncultured” or “s_”, with the latter indicating a lack of taxonomic resolution to identify the taxon at species level. As these generic proxies do not contribute with taxonomic information, these and other similar entries (namely, “uncultured”, “sp.”, “metagenome” and “human_gut”) were homogenized by replacing them with the lowest taxonomic rank with complete nomenclature and the corresponding taxon.

**Blood count data**

Complete blood count data spanning the first year after transplant was collected retrospectively from the blood test results database of our institution. Results from dates close to the oral sampling phases (aplasia not included due to the necessarily low counts for all patients) and 3, 6, 9, and 12 months after stem-cell infusion were collected. The median gap between these target periods and the actual blood test dates was 0 for all target periods. Outlier counts were identified by running the results collected altogether (independently of the period) through the Grubbs’ test and removed. Cell counts too low to be accurately quantified were set to 1.
